# Using quantitative magnetic resonance imaging to track cerebral alterations in multiple sclerosis brain: a longitudinal study

**DOI:** 10.1101/2022.01.26.22269806

**Authors:** Nora Vandeleene, Camille Guillemin, Solène Dauby, Florence Requier, Maëlle Charonitis, Daphne Chylinski, Evelyne Balteau, Pierre Maquet, Emilie Lommers, Christophe Phillips

## Abstract

Quantitative MRI quantifies tissue microstructural properties and supports the characterization of cerebral tissue damages. With an MPM protocol, 4 parameter maps are constructed: MTsat, PD, R1 and R2*, reflecting tissue physical properties associated with iron and myelin contents. Thus, qMRI is a good candidate for *in* vivo monitoring of cerebral damage and repair mechanisms related to MS. Here, we used qMRI to investigate the longitudinal microstructural changes in MS brain.

Seventeen MS patients (age 25-65, 11 RRMS) were scanned on a 3T MRI, in two sessions separated with a median of 30 months, and the parameters evolution was evaluated within several tissue classes: NAWM, NACGM and NADGM, as well as focal WM lesions. An individual annual rate of change for each qMRI parameter was computed, and its correlation to clinical status was evaluated. For WM plaques, three areas were defined, and a GLMM tested the effect of area, time points, and their interaction on each median qMRI parameter value.

Patients with a better clinical evolution, i.e., clinically stable or improving state, showed positive annual rate of change in MTsat and R2* within NAWM and NACGM, suggesting repair mechanisms in terms of increased myelin content and/or axonal density as well as edema/inflammation resorption. When examining WM lesions, qMRI parameters within surrounding NAWM showed microstructural modifications, even before any focal lesion is visible on conventional FLAIR MRI.

The results illustrate the benefit of multiple qMRI data in monitoring subtle changes within normal appearing brain tissues and plaque dynamics in relation with tissue repair or disease progression.

**Key points:**

- Patients with a better clinical evolution showed microstructural improvement in term of MTsat and R2* increase in their normal appearing tissue, suggesting repair mechanisms.
- Using qMRI allows to detect modifications in tissue microstructure in normal appearing tissues surrounding lesions several months before they are visible on conventional MRI.

## 1. Introduction

Multiple sclerosis (MS) is a chronic autoimmune disease of the central nervous system (CNS). Plaques are the pathological hallmark of MS. They are spread in acute, focal, disseminated and recurrent way throughout the CNS and harbor variable degrees of inflammation, demyelination, gliosis and axonal injury [1, 2]. Plaques are not restricted to the white matter (WM), but are also present in the cortex and deep grey matter (GM) [3-5].

Over and above focal WM lesions, an early, diffuse and chronic inflammation within the normal appearing white matter (NAWM) and grey matter (NAGM) is ultimately responsible for diffuse neuro-axonal loss and neurodegeneration, which is deemed responsible for a progressive accumulation of disability [3, 4, 6].

By contrast, effective repair mechanisms can occur within focal lesions but probably also in normal appearing brain tissue (NABT) [7]. However, our understanding of these complex processes is still fragmentary. The difficulty of acquiring histopathological data on MS patients at various stages of the disease makes it challenging to describe the time course of injury and potential repair mechanisms in MS. Consequently, there is a need for new imaging techniques to improve the *in-vivo* monitoring of brain damages formation, progression and repair in MS [8].

Conventional MRI (cMRI) readily depicts focal WM lesions on T2/FLAIR sequences and is able to distinguish between acute and allegedly chronic lesions. T2-hyperintensities in cMRI constitute the keystone of McDonald diagnostic criteria [9] and also make an important contribution to the monitoring of WM lesion burden. Unfortunately, cMRI sequences do not sensitively detect cortical lesions and diffuse changes in NABT, due to a rather low sensitivity of cMR imaging for cortical lesions, mixed contrast weight, and an overall limited histopathological specificity within cerebral tissues. Quantitative MRI (qMRI) potentially overcomes these limitations by quantifying physical microstructural properties of cerebral tissue in standardized units. qMRI is more sensitive but also more specific to microstructural properties of CNS tissues. Magnetization transfer ratio (MTR) was regularly linked to cerebral macromolecular content detected by a greater percentage loss of magnetization in voxels with a higher myelin content and axons density [10-12]. Post-mortem studies comparing the relative contribution of these two factors indicate that myelin has a stronger and more direct influence on MTR than the axonal density, which is considered as a T1-dependent effect. Tissue water content (inflammation, edema…), another T1-dependent effect, also accounts for MTR variability [11, 13, 14]. However, the MT saturation (MTsat) map offers a measure which, unlike MTR, is minimally affected by longitudinal relaxation and B1 mapping inhomogeneities [15], increasing its sensitivity to myelin content. Moreover, the brain contrast to noise ratio is larger for the MTsat map than for MTR, thus improving brain tissue segmentation in healthy subjects [11, 16]. R2* was usually linked to iron and myelin contents, as paramagnetic iron and diamagnetic myelin generate microscopic field gradients in the CNS, thus shortening T2* and increasing R2* (1/T2*). Orientation and density of myelin fibers are also a determining factor of R2* values [17-19]. Iron is probably a key factor in MS monitoring as it was shown that aberrant iron metabolism occurs in the course of the disease [20]. Particularly increased iron concentration within chronic active lesions (i.e iron rim lesion) or deep grey matter structures was observed [20]. Regarding the longitudinal relaxation rate R1 (1/T1), its three major determinants in the CNS are tissue myelination and associated axons, iron, and extracellular water contents [19, 21, 22]. Finally, proton density (PD) mostly reflects the free water content of the brain [23].

A number of cross-sectional studies using a combination of MT, R1, R2* or PD parameters, comparing MS patients to healthy controls, reported significant changes in the microstructure of NABT, such as a decrease in MT, R1 and R2* and an increase in PD in patients [24-32]. Few studies addresses the longitudinal variations in qMRI. R2* [33-35], PD and T1 were reported to increase in the basal ganglia over a period of a year [36], whereas a decrease in MTR in NAWM was reported over one [37] or two years [38].

Regarding focal WM plaques, qMRI emerges as an appealing biomarker to describe the dynamic processes of demyelination and remyelination. For instance, MTR was shown to sharply decrease within gadolinium enhancing lesions before recovering during the subsequent months [39-41], and within NAWM days to weeks before the formation of a new active lesion [42, 43].

Because each qMRI parameter is differently sensitive to histologically measured iron and myelin contents, this approach might become a fundamental tool for longitudinal *in vivo* monitoring of MS lesions and NABT evolution at the tissue microstructural level.

In this longitudinal study, we investigate the evolution of four simultaneously acquired qMRI parameters (MTsat, PD, R1, R2*) within NABT and WM lesions of 17 MS patients - relapsing remitting (RRMS) and progressive MS (PMS) - who were scanned two times with at least a one-year interval, following the same multi-parameter mapping (MPM) protocol at 3 Tesla [10, 44].

We assessed the time course of parameter values in several tissue classes: normal appearing white matter (NAWM), normal appearing cortical and deep GM (NACGM and NADGM) as well as focal WM lesions. In addition, we related longitudinal qMRI changes within NABT to clinical course.

## 2. Materials and methods

### 2.1 Population

Seventeen patients, recruited at the specialized MS outpatient clinic of the CHU Liège, Belgium, with a diagnosis of MS according to the McDonald criteria 2010 [45], were gathered from two studies: ten of them were part of the work reported by Lommers et al. 2019 [24], the other seven were recruited from another MS study taking place at the GIGA Cyclotron Research Centre – In Vivo Imaging (Liège, Belgium) [46]. For the first study (10 subjects), the inclusion criteria were (1) age between 18 and 65 years ; (2) Expanded Disability Status Scale (EDSS) inferior or equal to 6.5 ; (3) absence of relapse within the previous four weeks ; (4) absence of IVMP administration for at least 6 months prior to the study. Both RRMS and PMS patients were recruited. The second study (7 subjects) differs a bit as it comprises only RRMS patients, and the inclusion criteria were (1) age between 18 and 45, (2) EDSS between 0 and 4, (3) absence of relapse for at least 6 months prior to the study, (4) disease duration was below or equal to 5 years, (5) absence of IVMP administration for at least 6 months prior to the study. For both studies, compatibility with MRI and absence of other neurological/psychiatric diseases were required. These studies were approved by the local ethics committee (approval numbers B707201213806 and B707201835630, respectively). All patients were followed up and scanned twice on the same 3T MRI scanner, every 1 to 3 years. For each of the 17 MS patients, data from two MRI sessions were available, at T0 and T1. This cohort included 11 RRMS and 6 (primary and secondary) PMS patients. Thirteen were receiving disease-modifying treatments (DMTs). The patients’ median age was 36 years (range: 25-65) and the median time interval between two scans was 30 months (range: 14-61). Demographic data appears in Table 1. Extended individual information appears in Supplementary data.

**Table 1:** Demographic data of the study sample

|  |  |  |
| --- | --- | --- |
|  | *All Patients (n = 17)* |  |
| *Age, y, median (range)* | 36 (25-65) |  |
| *Sex, F/M* | 7/10 |  |
| *MS phenotype (RRMS/MS)* | 11/6 |  |
| *Baseline disease duration, y, median (range)* | 3.4 (0.3-28) |  |
| *Baseline EDSS, median (range)* | 2.5 (1-6.5) |  |
| *Baseline number of relapses, median (range)* | RRMS: 2 (1-5) | PMS: N/A |
| *Disease-modifying treatment* | RRMS:<br>First line, n: 5<br>Second line, n: 6 | PMS:<br>Ocrelizumab, n: 2<br>None, n: 4 |

### 2.2 MR image acquisition

MRI data were acquired on a 3T whole-body MRI-scanner (Magnetom Prisma, Siemens Medical Solutions, Erlangen, Germany). The whole-brain MRI acquisitions included a multi-parameter mapping protocol (MPM), from which one can simultaneously estimate (semi)quantitative maps of magnetization transfer saturation (MTsat), proton density (PD), transverse relaxation (R1) and effective longitudinal relaxation (R2*). This protocol arising from an international collaborative effort [10, 44], has already been used to study brain microstructure in various conditions including normal aging [9, 44, 47], brain tumor [48], Parkinson’s disease [49-51] as well as MS. It consists of three co-localized 3D multi-echo fast low angle shot (FLASH) acquisitions at 1mm^3^ resolution and two additional calibration sequences to correct for inhomogeneities in the RF transmit field [52, 53]. The FLASH datasets were acquired with predominantly PD, T1 and MT weighting, referred to in the following as PDw, T1w and MTw, at multiple echo times. All three had high bandwidth to minimize off-resonance and chemical shift artifacts. Volumes were acquired in 176 sagittal slices using a 256×224 voxel matrix. GRAPPA parallel imaging was combined with partial Fourier acquisition to speed up acquisition time to approximately 20 min. An additional FLAIR sequence was recorded with spatial resolution 1mm^3^ and TR/TE/TI=5000ms/516ms/1800ms. Extra B1 field mapping images (transmit B1+ and receive B1-fields) were also acquired to reduce spatial inhomogeneities related to B1 effect. This was essential for proper quantification of T1 (or R1=1/T1) in particular. Finally, B0 field mapping images, corresponding to both magnitude images and pre-subtracted phase image, were acquired for image distortions corrections. A summary of the acquisition parameters appears in Supplementary data.

Note that these MR sequences at 3 Tesla are not sensitive to cortical lesion as described in [54, 55] although a few lesions at the cortico-subcortical border were detected. Quantification of cortical parameters is thus confounded by voxels potentially located within cortical lesions.

### 2.3 MR image processing

All data processing was performed in Matlab (The MathWorks Inc., Natick, MA, USA) using SPM12 (www.fil.ion.ucl.ac.uk/spm) and three additional dedicated SPM extensions: the Lesion Segmentation Tool (LST) version 1.2.3 (www.statisticalmodelling.de/lst.html) [56], the “quantitative MRI and in vivo histology using MRI” toolbox (hMRI, http://hmri.info) [10], and “US-with-Lesion” tool (USwL, https://github.com/CyclotronResearchCentre/USwLesion).

Quantitative maps - MTsat, PD, R1 and R2* - were estimated using the hMRI toolbox. T1w, PDw and MTw images acquired at multiple echo times (TE) were extrapolated to TE=0 to increase signal-to-noise ratio and remove the otherwise remaining R2* bias [10, 24, 57]. The TE=0 extrapolated MTw, PDw and T1w images were used to calculate MT saturation, R1 and apparent signal amplitude A* maps. PD map was derived from A* map, which is proportional to proton density. All quantitative maps were corrected for inhomogeneities from local RF transmit field (B1+), and R1 quantitative maps were further corrected for imperfect RF spoiling using the strategy of Preibisch and Deichmann [58]. The receive bias field map (B1-) was used to correct PD maps for instrumental biases. The R2* map was estimated from all three multi-echo series (MTw, PDw and R1w) using the ESTATICS model [57].

After generating quantitative maps using the hMRI toolbox for all sessions, spatial preprocessing involved the following steps (Figure 1): within-patient registration brought the two serial MR data sets into the individual T0 space, using the longitudinal registration tool from SPM [59]. For each individual patient, a preliminary WM lesion mask was generated based on FLAIR and T1w images by the lesion growth algorithm implemented in the LST toolbox [56], followed by manual corrections by an MS expert (EL) to remove aberrant/artefactual lesion detections [24]. The images were then segmented using the USwL toolbox, which consists of an extended version of the traditional Unified Segmentation (US) algorithm [60] and includes an additional tissue class representing the WM lesion(s). The US-with-lesion method internally generates a subject-specific extended set of tissue probability maps (TPM) [61]: an extra tissue class, based on the smoothed preliminary lesion mask warped into template space (using cost function masking during normalization [62]), is added to account for the lesion, and the original white matter prior map is updated accordingly [63]. The grey matter TPM was not updated due to a very low number of lesions present in the cortical ribbon. Multi-channel segmentation was conducted, using MTsat, PD, R1 and FLAIR images. This pipeline did not use the PD-, T1- and MT-weighted images acquired for the MPM maps construction, but the parametric maps themselves instead. In this way, voxels do not depict MR intensities but rather physical quantitative parameters. The method generated the segmented tissue classes (*a posteriori* tissue, including lesion, probability maps), as well as spatial warping into standard template space. The preliminary lesion mask was used as input for the first session data (at T0) then the *a posteriori* lesion map generated at this initial step served as prior to the subsequent session (at T1).

**Figure 1:**
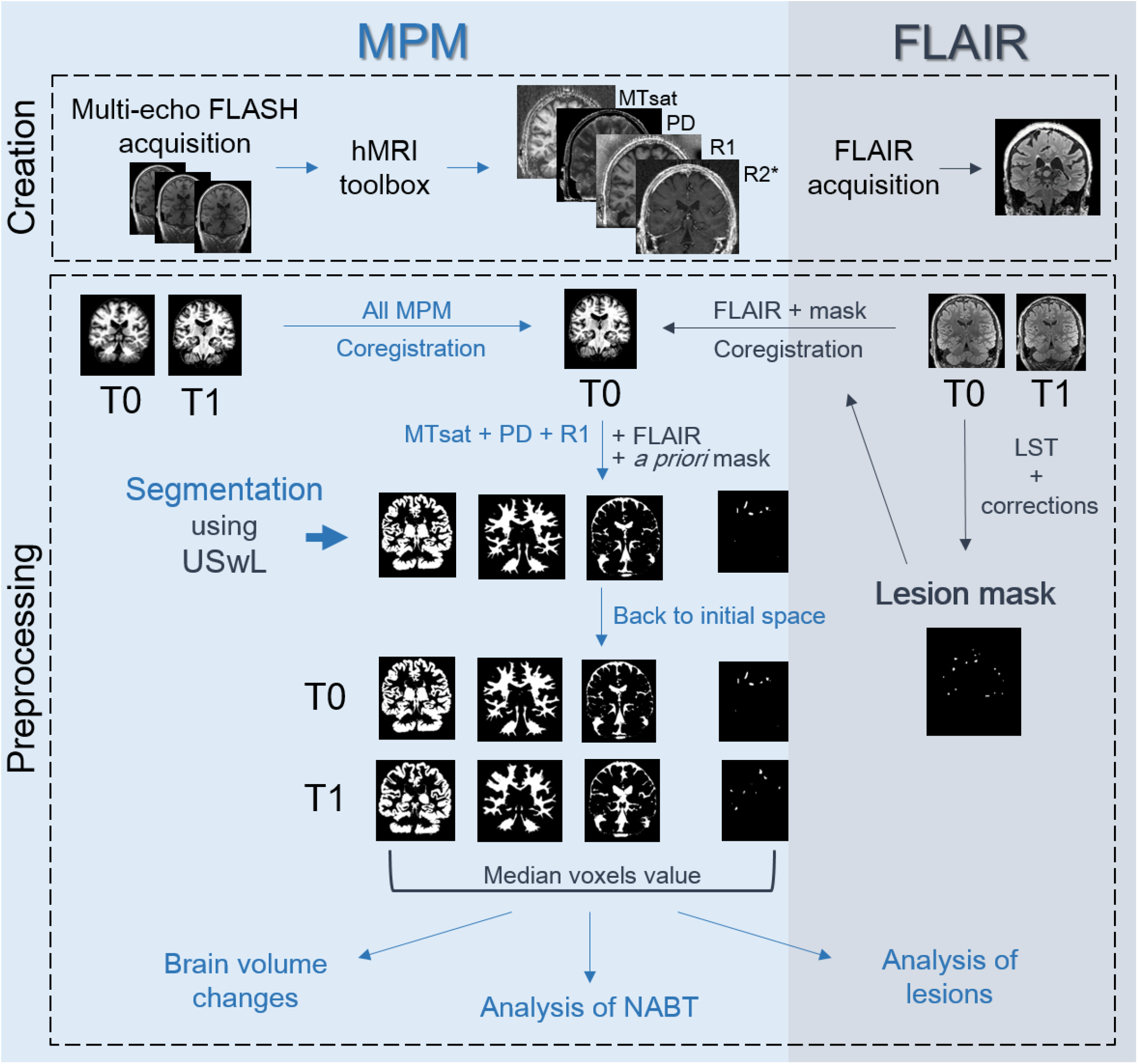
Chartflow of data creation and processing (see text). MPM maps were created with the hMRI-toolbox, FLAIR images were directly acquired for both sessions (T0 and T1). A preliminary mask was constructed based on T0 FLAIR. All images (MPM and FLAIR, T0 and T1) were co-registered to the MPM T0 space. Segmentation using USwL allowed to isolate the different tissue classes.

Segmentation teased out the different tissue classes of interest: NAWM, NACGM and NADGM, as well as WM lesions. To analyze the microstructure within those tissue classes, *a posteriori* tissue maps were binarized and tissue-specific independent masks were constructed: each voxel is assigned to one single tissue class with the highest probability for that voxel (provided that this probability was above 0.2). The lesion binary mask was further cleaned for lesions <10mm^3^ which likely resulted from segmentation errors. Finally, binarized tissue class masks were in turn applied on the MPM maps to extract voxel values inside them.

### 2.4 Brain volume change

Volumetric changes were investigated using the USwL *a posteriori* tissue probability maps. The following measures of brain volume were computed for each session of each participant: (1) Total intra-cranial volume (TIV) = volume (NAWM + GM + CSF + lesions), (2) brain parenchymal fraction (BPF) = volume (NAWM + GM + lesions)/TIV, (3) Gray matter fraction (GMF) = volume (GM)/TIV, and (4) lesion fraction (LF) = volume (lesion)/TIV. The percentage of change between both scanning sessions was evaluated for each volumetric measurement, then annualized changes were computed by dividing these measures by scan intervals (in years). Results were directly analysed with a t-test (testing if significantly different from 0 at *p* < .05), but also in the same way as the normal appearing tissues MR parameters in relation to the patients’ clinical status (see next section).

### 2.5 Analysis of normal appearing tissues

The median value of quantitative MRI parameters was extracted from the three normal appearing tissues (NAWM, NACGM and NADGM), and an individual annual rate of change (ARoC) was computed for each parameter in each tissue class, based on the initial and final values and accounting for the time interval (in years) between scans. This rate of change in qMRI parameters served as dependent variable in a general linear model testing the effect of clinical status:

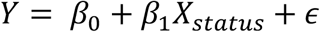

*Y* is the ARoC for a qMRI parameter and tissue class, *β*’s are the regression parameters corresponding to the associated regressor (with *β*_0_ the intercept), and *ϵ* the residuals. *X*_*status*_ is a binary categorical variable representing the patient’s disease activity status: a status score of 1 was assigned to patients stable or improving from T0 to T1.

This patient status *X*_*status*_ was derived from one score of disease activity: NEDA-3 (No Evidence of Disease Activity [64]), a composite of three related measures of disease activity. A score of 0 was assigned in the presence of new clinical relapses (only concerning RRMS patients) and/or MRI activity (new or enlarged lesions visible on FLAIR T2 or Gadolinium-enhanced images) and/or sustained disability progression over six months based on Expanded Disability Status Scale (EDSS). For both RRMS and PMS patients, disability progression was defined as a 1.0-point increase if the EDSS score was ≤ 4.0 at baseline and as a 0.5 point increased if the baseline EDSS score was > 4.0. The threshold of 4.0 was proposed in this study because it is considered as a milestone regarding ambulatory performance.

NEDA-3 and was evaluated at mid- and end-scanning interval, and a final status score of 0 was given only to patients for which disease activity or progression was noted in both cases, indicating a clear progression of the disease over the whole interscan interval.

The influence of several clinical measurements such as 25 FWT, 9HPT and SDMT was also considered to refine the evaluation of disease activity. However complete data were lacking for several patients. Moreover, when available, these additional clinical parameters did not modify the final *X*_*status*_. Longitudinal clinical information allowing to derive the disease activity status for each subject appears in Table 2. Additional clinical information concerning annual relapse rate and treatment administration appears in Supplementary material.

**Table 2:** Longitudinal clinical information and derived disease status score. The time period between T0 and T1 is expressed in months.

|  | EDSS<br>T0 | EDSS<br>T1/2 | New<br>lesion<br>T1/2 | Relapse<br>T1/2 | NEDA<br>T1/2 | EDSS<br>T1 | New<br>lesion<br>T1 | Relapse<br>T1 | NEDA<br>T1 | Time<br>period<br>T0-T1 | Score |
| --- | --- | --- | --- | --- | --- | --- | --- | --- | --- | --- | --- |
| sub-001 | 2 | 2 | None | None | YES | 2 | None | None | YES | 30 | 1 |
| sub-002 | 1.5 | 1.5 | None | None | YES | 1.5 | None | None | YES | 27 | 1 |
| sub-003 | 2 | 2 | None | None | YES | 2 | None | None | YES | 27 | 1 |
| sub-004 | 3 | 3 | None | None | YES | 3.5 | None | None | YES | 25 | 1 |
| sub-005 | 1 | 1 | None | None | YES | 1 | None | None | YES | 24 | 1 |
| sub-006 | 1.5 | 1.5 | None | None | YES | 1.5 | None | None | YES | 24 | 1 |
| sub-007 | 2 | 2 | None | None | YES | 2 | None | None | YES | 22 | 1 |
| sub-008 | 3.5 | 4.5 | None | N/A | NO | 5 | None | N/A | NO | 51 | 0 |
| sub-009 | 2 | 2.5 | None | None | YES | 2.5 | None | None | YES | 57 | 1 |
| sub-010 | 6 | 6 | Yes | N/A | NO | 6.5 | None | None | NO | 14 | 0 |
| sub-011 | 6 | 6 | None | N/A | YES | 6.5 | None | N/A | NO | 14 | 1 |
| sub-012 | 1 | 1.5 | None | None | YES | 1.5 | None | None | YES | 55 | 1 |
| sub-013 | 5.5 | 6 | None | N/A | NO | 6.5 | None | N/A | NO | 60 | 0 |
| sub-014 | 2.5 | 3.5 | Yes | Yes | NO | 3 | None | None | YES | 57 | 1 |
| sub-015 | 4 | 4.5 | None | N/A | NO | 5 | None | N/A | NO | 51 | 0 |
| sub-016 | 5 | 4.5 | Yes | N/A | NO | 4.5 | None | N/A | YES | 61 | 1 |
| sub-017 | 2 | 3 | Yes | Yes | NO | 3 | Yes | Yes | NO | 56 | 0 |

Permutation tests were employed for inferences [65]. R-squared value was tested against computed statistics after permutation of the data. For a number *n* of permutations, the *X*_*status*_ values were randomly shuffled (constructing a new regressor written 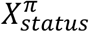), tested against the unchanged response *Y*, and generating each time a permuted R-squared value (noted *R*_*π*_, *R*_*obs*_ being the true R-squared value computed without permutation of the data).

The condition 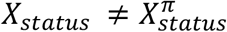 is verified at each permutation. After *n* permutations (with *n = 5000* in this study), a *p*-value was computed based on the following formula:

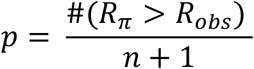

which estimates the probability of obtaining *R*_*obs*_ under the null hypothesis that *Y* is not correlated to *X*_*status*_. The null hypothesis is rejected if *p* < .05 FDR-corrected for multiple comparisons [66], for the 12 tests performed (3 tissue classes with 4 qMRI parameters).

Two-tailed t-tests were applied *post-hoc* on the significant results of permutation tests to compare the ARoC distribution between disease status, i.e., *X*_*status*_ = 0 against *X*_*status*_ = 1. Inferences were conducted at a significance level of .05.

The same pipeline was applied to the brain volumetric changes (BPF, GMF and LF) to test their correlation to the disease activity status.

### 2.6 Analysis of lesions and peripheral tissues

For white matter lesions analysis, we did not use ARoC but exploited directly the qMRI parameters voxel values. Importantly, with USwL segmentation, the prior lesion mask is only used in a probabilistic way and the estimated posterior lesion map, obtained using MTsat, PD, R1 and FLAIR images, typically showed more extended lesion than clinically visible on the FLAIR image alone. Therefore, we separated focal lesions detected on FLAIR images, with LST segmentation and visual inspection, from their peripheral regions detected on qMRI maps. Two different peripheral regions were considered: one for each time point (T0 and T1). Therefore, at T0, three distinct lesion-related regions were isolated:

- The lesions, as clinically defined, pertaining to hyperintensity on the conventional FLAIR MR image acquired at T0. These are referred to as ‘focal FLAIR lesion’.
- The peripheral region detected on qMRI maps at T0, at the borders of (but not including) the focal FLAIR lesion. Those are referred to as ‘initial peripheral lesion’.
- The peripheral region, detected on qMRI maps at follow up, bordering (but not including) the initial peripheral lesion, further referred to as ‘later peripheral lesion’. This was computed by masking out the T1 lesion mask with the T0 lesion mask. This region allows us to determine whether its microstructure at T0 forebodes a full-blown plaque, detectable during follow up. Those sometimes appear hyperintense on FLAIR images.

The three areas were compared between each other and with NAWM, in order to characterize them on a microstructural basis (Figure 2). For an accurate lesion-by-lesion analysis, only enlarging lesions, i.e., present in the three masks, were considered for these comparisons. NAWM region consisted of all white matter voxels which did not belong to any of the three lesion-related regions. The four areas are not overlapping as no voxel could belong to more than one class at the same time.

**Figure 2:**
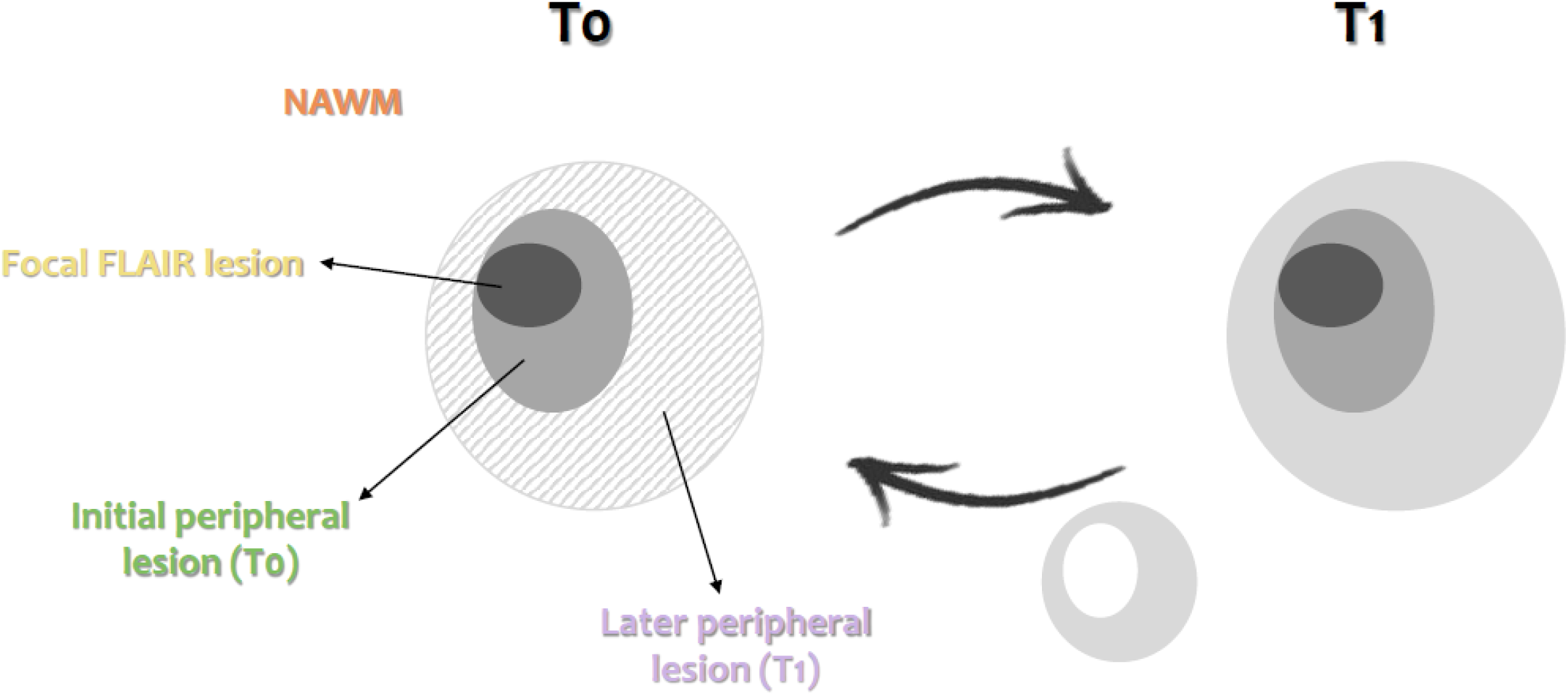
Schematic illustration of the NAWM and 3 lesions-related areas: focal FLAIR lesion (dark gray area), initial peripheral lesion detected at T0 (medium gray area), later peripheral lesion detected at T1 (dashed, left, and light gray, right, area).

For all participants, MTsat, PD, R1 and R2* median values were extracted from each lesion area, considering lesions individually (between 2 and 66 measurements per subject). Similarly, the median qMRI values within NAWM were also extracted (one measurement per subject). These values were extracted from T0 and T1 scans separately. Statistical analyses were performed in SAS 9.4 (SAS Institute, Cary, NC). None of the qMRI parameter was normally distributed, therefore we applied a log transformation on each of them prior to statistical analysis. For each qMRI parameter, a separate Generalized Linear Mixed Model (GLMM) tested the effect of areas (NAWM and the three lesion-related areas), and time points (T0 and T1), as well as their interaction (i.e., area*time), on the median qMRI parameter value, with a first-order autoregressive variance/covariance model and participants as a random factor (intercept). The degrees of freedom were estimated using Kenward-Roger’s method. Statistical significance was estimated at *p* < .05 after adjustment for multiple comparison using Tukey’s procedure.

## 3. Results

### 3.1 Volume changes

Brain parenchymal fraction (BPF) annually decreased between T0 and T1 by -0.67 ± 1.12% (significantly different from zero; paired-sample t-tests; *t*(16) = 2.57; *p* = .0204) whereas lesion fraction (LF) increased by 22.88 ± 26.13% (*t*(16) = −3.70; *p* = .0019). GM fraction (GMF) non-significantly decreased by -0.30 ± 1.44%.

### 3.2 Analysis of normal appearing tissues

As expected, changes in MTsat and R2* within normal appearing brain tissues (NABT) between T0 and T1 varied across subjects (Figure 3). PD and R1 exhibited similar behaviors, see Supplementary data.

**Figure 3:**
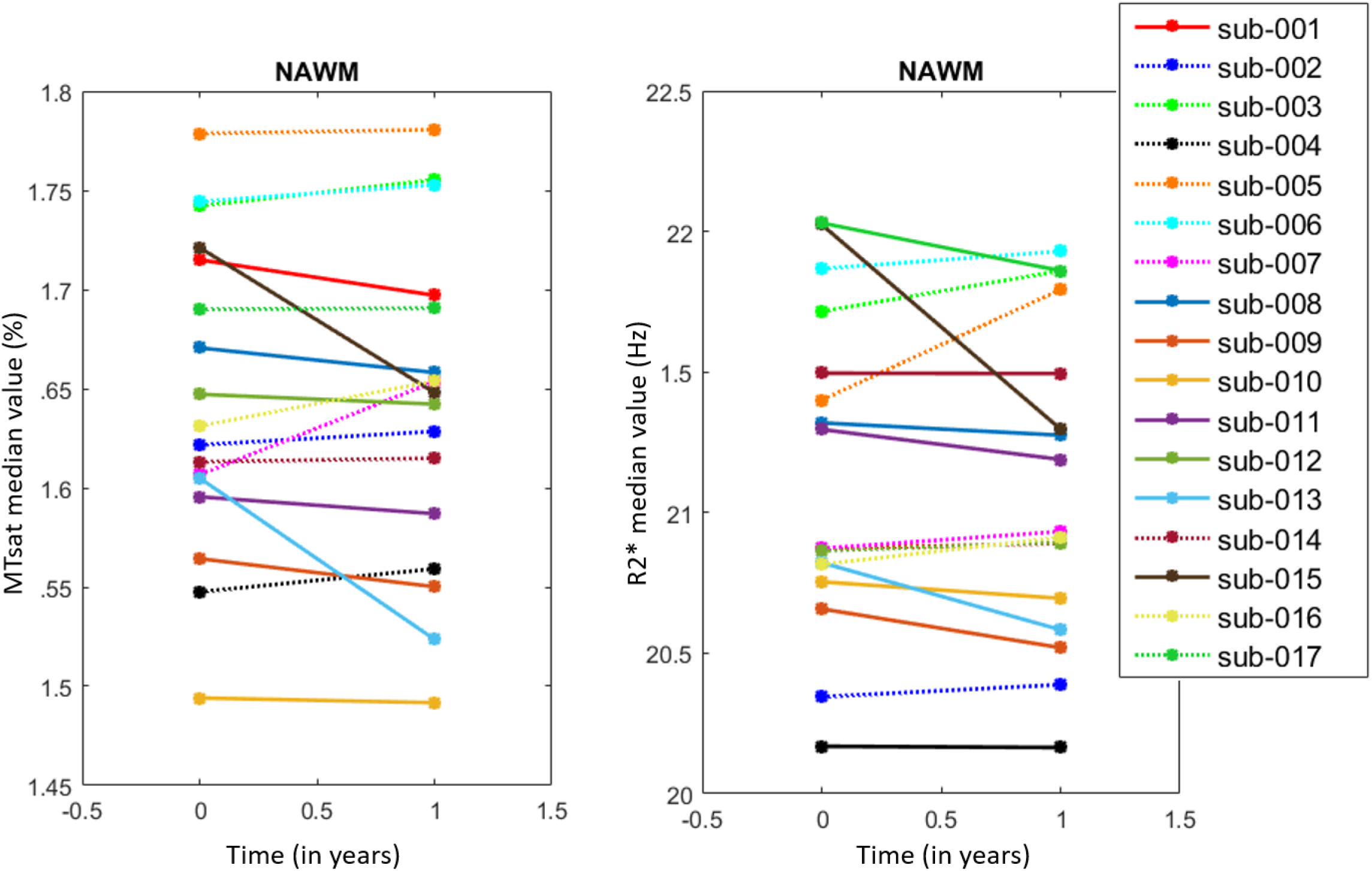
Line plots illustrating individual ARoC’s for MTsat (left) and R2* (right) in NAWM. Each line corresponds to one subject. Dotted lines represent increasing rates.

At the group level, with the regression analysis and permutation inference, we observed that the annual rate of change (ARoC) of MTsat and R2* positively regressed with disease status as follows (Table 3): MTsat in NAWM and NACGM and R2* in NAWM significantly increased in patients who fare well (*X*_*status*_ = 1).

**Table 3:** Regression coefficients and their associated *p*-values (in parentheses) for the effects of *X*_*status*_ on the individual ARoC for each qMRI parameter (MTsat, PD, R1 and R2*) and for volumetric measurements (BPF and LF). * Results significant at *p* < .05, FDR corrected.

|  | NAWM | NACGM | NADGM |
| --- | --- | --- | --- |
| **MTsat** | **0.039 (.011)\*** | **0.017 (.007)\*** | 0.004 (.749) |
| **PD** | -0.018 (.670) | 0.405 (.225) | 0.250 (.552) |
| **R1** | 0.009 (.139) | 0.004 (.471) | 0.010 (.111) |
| **R2\*** | **0.295 (.002)\*** | 0.121 (.092) | 0.066 (.770) |
| **BPF** | -0.884 (.1562) |  |  |
| **LF** | 21.23 (.1082) |  |  |

*Post-hoc* t-tests applied on these significant results for a clearer illustration of the difference in disease status (Figure 4) were all significant at a level of .05.

**Figure 4:**
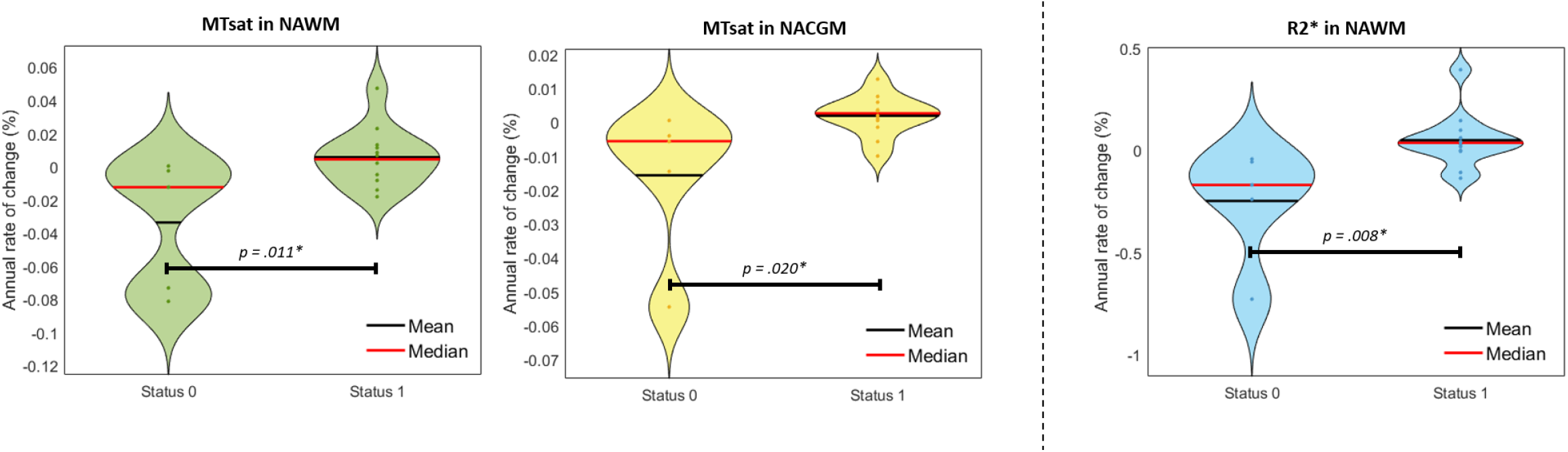
Violin plots of significant change rates in microstructure with respect to *X*_*status*_. From left to right: MTsat in NAWM, MTsat in NACGM, R2* in NAWM. *\* P* < .05.

Regarding BPF and LF, their correlation to the disease activity status was not significant (Table 3), suggesting that qMRI parameters are more sensitive to subtle microstructural changes in normal appearing tissues over time than global morphological measurements

### 3.3 Analysis of lesion microstructure

The number of enlarging WM lesions between T0 and T1 varied from 2 to 66 across patients, for a total of 741 identified enlarging lesions among all subject, corresponding on average among patients to 63% (±31%) of the amount of initial focal lesions. The number of enlarging lesions did not significantly differ between patients’ disease status groups (*t*(15) = .244, *p* = .811).

GLMMs found a significant effect of areas (3 lesion regions and NAWM) for MTsat, R1, R2* and PD median (MTsat: *F*_3_ = 35.34, *p* < .0001, PD: *F*_3_ = 68.03, *p* < .0001, R1: *F*_3_ = 40.26, *p* < .0001, R2*: *F*_3_ = 32.32, *p* < .0001). By contrast, neither time effect (T0 vs T1; MTsat: *F*_3_ = 0.36, *p* = .5481, PD: *F*_3_ = 1.20, *p* = .2735, R1: *F*_3_ = 2.05, *p* = .1520, R2*: *F*_3_ = 2.86, *p* = .0911), nor the area*time interaction (MTsat: *F*_3_ = 0.09, *p* = .9671, PD: *F*_3_ = 0.14, *p* = .9346, R1: *F*_3_ = 0.14, *p* = .9331, R2*: *F*_3_ = 0.40, *p* = .7565) were significant, suggesting the microstructural stability of the initial lesion core. *Post-hoc* tests confirmed significant differences between the four tissue areas.

At times T0 and T1, MTsat, R1 and R2* values were significantly larger in the initial peripheral lesion than FLAIR lesion, in the later peripheral lesion than the initial one, and in the NAWM than later peripheral lesion. The reverse was observed for PD. The significant difference in parameters between initial and later peripheral lesion at T0 suggests that subtle microstructural changes appear in the periphery of the initial lesion, months before their detection as focal FLAIR lesions at T1. Adjusted *p*-values appear in Figure 5. Detailed statistical results of the GLMM’s appear in Supplementary data.

**Figure 5:**
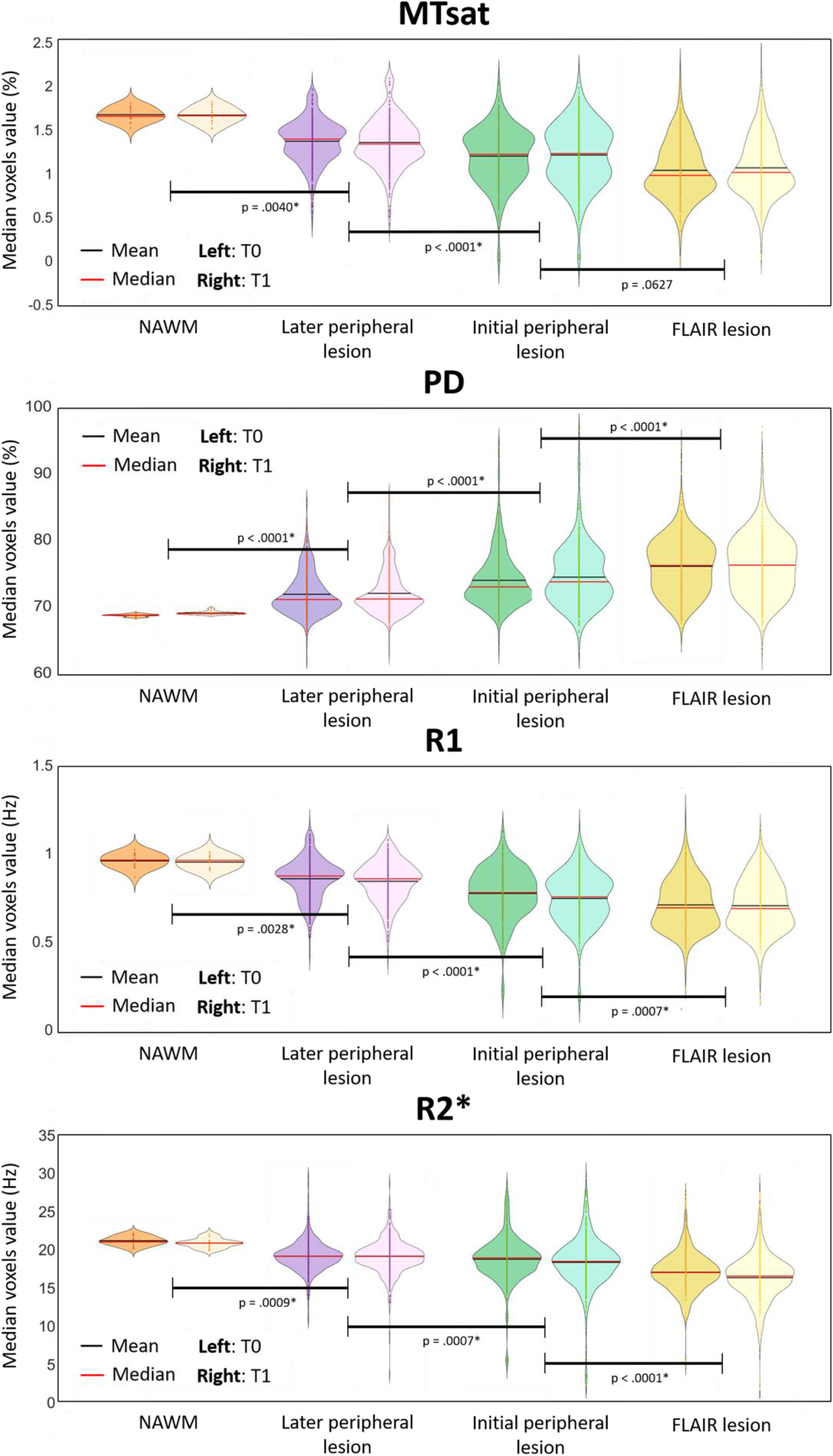
Microstructural parameters in NAWM and the 3 lesion-related areas, for each scanning time T0 and T1. P-values were obtained with *post-hoc* tests on the tissue area effect. * *P* < .05.

## 4. Discussion

This longitudinal study followed up volumetric data and qMRI brain metrics (MTsat, PD, R1, R2*) in 17 patients with multiple sclerosis for a median time interval of 30 months. The main results are threefold. First, the microstructure of normal appearing brain tissues changes over time and these modifications concur with, and potentially drive, clinical evolution. This critical finding suggests that repair mechanism and edema resorption can be monitored *in vivo*. Second, the microstructure within WM plaques is remarkably heterogeneous. Importantly, at their periphery, microstructural alterations foreshadow their expansion, as detected by conventional MRI. Third, as expected, we observed a small but significant brain atrophy and lesion load increase with time.

### Quantitative MRI parameter time course within NABT

In this study, we used a multiparameter mapping protocol that was gradually optimized and validated for multi-centric studies [67]. It provides high-resolution maps of multiple qMRI parameters from data acquired during a single scanning session of acceptable duration. A number of cross-sectional studies using a combination of MT, R1, R2* or PD parameters reported significant changes in the microstructure of NABT in MS [24-32]. By contrast, longitudinal analyses of multiparameter qMRI data are scarce. A progressive shortening of T2/T2* [68] or increase in R2* [33-35] was reported within the basal ganglia, suggesting increased of myelin and/or iron contents as well as edema resorption. Likewise, PD and T1 increased within a year, suggesting a demyelination and/or axonal loss [36]. MTR progressively decreases in NAWM of MS patients over one [37] or two years [38]. These abnormalities tend to be more pronounced in progressive phenotypes [69] and were associated to a slow, diffuse and global myelin pathology.

Here, we showed that MTsat within NAWM and NACGM and R2* values within NAWM increase in clinically stable or improving patients. Because MTsat and R2* both correlate with myelin content [11, 47, 70-73], our results suggest repair mechanisms within NABT of patients who are responding to disease modifying treatments, despite the initial myelin/axonal loss and independently from WM focal lesion evolution. Such increases could also be explained by an edema/inflammation resorption, but less likely than myelin/axonal density changes since MTsat is the least dependent to water content among the four qMRI parameters. These results echo cross-sectional analyses showing that healthy controls (HC) have higher MTsat and R2* values within the same tissue classes compared to MS patients [24]. Annual rates of change of R1 and PD within NABT were not significantly associated with the individual clinical status in this study, although R1 reduction within NABT has already been reported in cross sectional [24, 29, 30] and longitudinal [36] studies comparing MS subjects to HC.

### Lesion microstructure

Focal inflammatory demyelinating lesions have been extensively characterized and are traditionally classified as active, chronic active (smoldering) or inactive plaques according to the presence and distribution of plaque-infiltrating macrophages/microglia [74-76]]. Focal WM pathology is a constantly evolving process including episodes of demyelination and remyelination but also accumulation of irreversible axonal damage. Age, disease duration, clinical phenotype as well as disease modifying treatment all contribute to the dynamic nature of focal WM pathology [75, 77]. This accounts for the large inter- and intra-individual heterogeneity of MS, which conventional MRI is largely unable to capture. By contrast, quantitative MRI parameters are sensitive to myelin, axonal as well as iron contents and appear as promising markers of plaque dynamics. For instance, MTR was shown to sharply decrease within gadolinium enhancing lesions before recovering during the subsequent months [39-41]. Likewise, reduction of MTR within NAWM, days to weeks before the formation of a new active lesion was also demonstrated [42, 43], and long-term MTR changes in WM plaques were observed in relation with disease progression [69, 78]. The present study broadens the quantitative characterization of plaque dynamics, in keeping with previous longitudinal studies [68, 79]. Two important findings emerge from the results. First, qMRI refines lesion segmentation, as compared to the processing based on the sole FLAIR image. In consequence, the initial lesion revealed by qMRI is typically wider that the plaque detected in FLAIR. Its periphery is characterized by a decrease in MTsat and R2* as compared to NAWM, suggesting an incipient demyelination, reminiscent of the so-called ‘periplaques’ [80]. Moreover, MTsat, R2* and R1 values progressively decrease from NAWM to plaque core, suggesting a centripetal loss of myelin content. Second, plaque microstructure is altered in plaque periphery before any observable change in conventional MRI signals. This finding suggests, in keeping with neuropathological observations [75, 77, 81, 82] that subclinical ongoing inflammation and/or demyelination takes place in the periphery of an active plaque, well before it is detectable on FLAIR or T1 post-gadolinium sequences. If confirmed on larger population samples, this finding might significantly modify treatment management in MS patients.

Another plausible hypothesis explaining the progressive decrease of R2* in initial and later peripheral regions is that iron-containing macrophages could be removing iron from the lesions through perivascular drainage into the extracellular compartment. Previous neuropathological studies have reported an iron loss at the edges of a subset of MS lesions, depending on their type (active, inactive, smoldering, etc.) as well as the patient’s age and disease duration [83, 84]. Due to the limited sensitivity of R2* to local iron concentration as compared, for example, to the combined use of R2* and quantitative susceptibility mapping (QSM) [18], validating this theory would require additional measures which can better describe iron dynamics in MS lesions and NAWM.

### Volumetric Data

CNS atrophy occurs in all stages of MS, since the preclinical phase of the disease and progresses throughout its course, at a much higher rate than one reported in normal aging [85-88]. In this study, the annual brain percentage volume loss at the group level was 0.67%, which is in line with previous publications [89]. We also showed a significant increase in lesion fraction. Volumetric data (ARoC’s) were highly variable across subjects: changes in BPF range from -2.52 to 1.17% and LF from -0.78 to 103.06%. This variability arises from a large number of factors which do not necessarily relate to MS: age, disease duration, disease phenotype, disease modifying treatment, circadian rhythm, hydration… [87, 88]. Moreover, annual changes in brain parenchymal fraction as well as lesion fraction only partially correlated to patients’ disease status, in accordance with a large amount of publications [36, 90]. This highlights the lack of specificity and sensitivity of volumetric measurements, at least at the individual level.

It can appear odd that brain atrophy progresses in parallel to repair mechanisms, as suggested by qMRI parameters. However, BPF reduction is minimal, and is not significant (see Table 3) between T0 and T1. One should keep in mind that cortical atrophy is an irreversible phenomenon. Given the inter- and intra-individual heterogeneity of MS progression, it is possible that patients who have undergone neuron-axonal loss at some point in the disease might be able to remyelinize their remaining axons, hopefully through therapeutic intervention or lifestyle changes. Besides, axonal remyelination is not always effective. Here we showed that variations in MTsat and R2* correlated to the disease activity status, but our clinical evaluation based on EDSS is undoubtedly imprecise. Once again, the size and heterogeneity of our cohort limits the interpretation of such results.

### Study limitation

As mentioned here above, the small size and heterogeneous aspect of the present dataset constitute major limitations of this study. Indeed, it is composed of only 17 patients, with a rather broad range of characteristics such as age, disease duration, disease phenotype, disease modifying treatment, etc., which are known to influence the disability state of the patient and thus their ability to put together repair mechanisms within cerebral tissues [1, 75-77, 91, 92]. In addition, the time interval between two scanning sessions varied rather widely across patients (between 14 and 61 months), although it was brought back to an annual rate where possible. All of these parameters were imposed by standard clinical follow up. Therefore, these results should not be over-interpreted but are nevertheless promising and call for a replication with a larger and more homogeneous or controlled set of MS patients. Larger longitudinal studies are currently being held and will probably confirm these preliminary results.

A second limitation is the absence of longitudinal MRI data acquired in a control group of healthy subjects. However, we considered that literature of longitudinal studies of healthy subjects that analysed tissue microstructure could constitute a solution for comparison with MS patients. For example, in Bonnier et al. (2017) [68], the control group did not show any significant differences regarding T1, T2* or MTR measurements over two years, and the median age of their group is quite similar to ours (34,3 vs 36 years). Also, in Elkady et al. (2018) [33], they found no longitudinal R2* effect in their control groups, even with an age range superior to ours. Moreover, the median age of our population (< 60 years), as well as the short period between two scanning sessions (median of 14 months), suggests that microstructural alterations would not be noticeable in a healthy participants group, as many quantitative ageing studies detected differences over much larger time periods [44, 70, 93].

## 5. Conclusion

These preliminary results highlight the relevance of multiple qMRI data in the monitoring of MS disease, highlighting subtle changes within NABT and plaque dynamics in relation with repair or disease progression. Of course, large scale longitudinal study would be needed to reproduce these findings and better exploit the full potential of qMRI parameters.

## Supporting information

All supplemental files

## Data Availability

All data produced in the present study are available upon reasonable request to the authors

## Acknowledgements

The authors are particularly thankful for the patients who took part in this study. N.V., E.L. and C.P. are supported by the Fonds de la Recherche Scientifique (F.R.S-FNRS Belgium).

## Abbreviations

ARoC: Annual Rate of Change
BPF: Brain Parenchymal Fraction
CDP: Confirmed Disability Progression
cMRI: Conventional Magnetic Resonance Imaging
CNS: Central Nervous System
GMF: Grey Matter Fraction
GLMM: General Linear Mixed Model
LF: Lesion Fraction
MPM: Multi-Parameter Mapping
MRI: Magnetic Resonance Imaging
MS: Multiple Sclerosis
MT: Magnetization Transfer
MTR: Magnetization Transfer Ratio
MTsat: Saturated Magnetization Transfer
NABT: Normal Appearing Brain Tissue
NACGM: Normal Appearing Cortical Grey Matter
NADGM: Normal Appearing Deep Grey matter
NAWM: Normal Appearing White Matter
NEDA-3: No Evidence of Disease Activity
RRMS: Relapsing-Remitting Multiple Sclerosis
PD: Proton Density
PMS: Progressive Multiple Sclerosis
qMRI: Quantitative Magnetic Resonance Imaging
R1: Longitudinal Relaxation Rate (1/T1)
R2*: Transverse Relaxation Rate (1/T2*)
TIV: Total Intracranial Volume
TPM: Tissue Probability Map
US: Unified Segmentation
USwL: Unified Segmentation with Lesion

## Appendix A: Supplementary data legends

**Supplementary data 1:** Multi-echo 3D FLASH acquisition parameters for Siemens Magnetom PRISMA MRI

**Supplementary data 2:** Extended demographic data. Age, disease duration, EDSS and relapses values were taken at baseline.

**Supplementary data 3:** Additional follow-up clinical data for each subject.

**Supplementary data 4**: Line plots illustrating individual ARoC’s for PD (left) and R1 (right) in NAWM. Each line corresponds to one subject. Dotted lines represent increasing rates.

**Supplementary data 5:** Differences of lesion class Least Squares Means. First two columns correspond to tissue class labels (0 = NAWM, 1 = Later peripheral lesion, 2 = Initial peripheral lesion, 3 = FLAIR lesion).

## Notes

**Conflict interests** None.

### Competing Interest Statement

The authors have declared no competing interest.

### Funding Statement

The main contributors are supported by the Fonds de la Recherche Scientifique (F.R.S-FNRS Belgium)

### Author Declarations

Ethics committee/IRB of the University of Liege Hospital gave ethical approval for this work

### Summary of Updates

This version of the manuscript was based on the Reviewer's comments after submission to 'Brain and Behavior' The abstract and introduction were modified to better highlight the scientific interest of the method. Table 2 was added to the main text (initially in Supplementary material).

