## Supplementary material for "Using quantitative magnetic resonance imaging to track cerebral alterations in multiple sclerosis brain: a longitudinal study": All supplemental files

**Supplementary data 1:** Multi-echo 3D FLASH acquisition parameters for Siemens Magnetom PRISMA MRI

| | PDw | T1w | MTw |
|---|---|---|---|
| **TR [ms]** | 24.5 | 24.5 | 24.5 |
| **Flip angle [°]** | 6 | 21 | 6 |
| **Bipolar gradient echoes/TE [ms]** | 8/TE 2.34 – 18.72 | 8/TE 2.34 – 18.72 | 6/TE 2.34 – 14.04 |
| **Off-resonance Gaussian MT pulse** | N/A | N/A | FA: 220° Frequency offset: 2 [kHz] |
| **Bandwidth [Hz/Px]** | 465 | 465 | 465 |

**Supplementary data 2:** Extended demographic data. Age, disease duration, EDSS and relapses values were taken at baseline.

| | Age range | Sex | Disease duration | MS type | EDSS | Disease modifying treatment | Total number of relapses at T0 (inclusion) |
|---|---|---|---|---|---|---|---|
| **sub-001** | 36-40 | F | 0.8 | RRMS | 2 | First line | 1 |
| **sub-002** | 26-30 | F | 0.7 | RRMS | 1.5 | Second line | 2 |
| **sub-003** | 31-35 | M | 1.6 | RRMS | 2 | First line | 1 |
| **sub-004** | 26-30 | M | 1.8 | RRMS | 3 | Second line | 2 |
| **sub-005** | 36-40 | F | 3.4 | RRMS | 1 | Second line | 1 |
| **sub-006** | 21-25 | M | 0.3 | RRMS | 1.5 | First line | 1 |
| **sub-007** | 31-35 | F | 1.6 | RRMS | 2 | Second line | 2 |
| **sub-008** | 61-65 | M | 16 | PMS | 4 | None | N/A |
| **sub-009** | 31-35 | M | 11.4 | RRMS | 3 | Second line | 5 |
| **sub-010** | 31-35 | M | 10 | PMS | 6 | None | N/A |
| **sub-011** | 61-65 | M | 25 | PMS | 6 | None | N/A |
| **sub-012** | 26-30 | F | 4 | RRMS | 1 | First line | 1 |
| **sub-013** | 61-65 | F | 23.7 | PMS | 5.5 | None | N/A |
| **sub-014** | 51-55 | F | 28 | RRMS | 2.5 | First line | 4 |
| **sub-015** | 46-50 | M | 8.9 | PMS | 4 | Ocrelizumab | N/A |
| **sub-016** | 36-40 | M | 2 | PMS | 5 | Ocrelizumab | N/A |
| **sub-017** | 46-50 | M | 0.5 | RRMS | 2 | Second line | 2 |

**Supplementary data 3:** Additional follow-up clinical data for each subject.

| | Annual relapse rate | IV steroids | Treatment change |
|---|---|---|---|
| **sub-001** | 0 | None | None |
| **sub-002** | 0 | None | None |
| **sub-003** | 0 | None | None |
| **sub-004** | 0 | None | None |
| **sub-005** | 0 | None | None |
| **sub-006** | 0 | None | None |
| **sub-007** | 0 | None | Switch from natalizumab to ocrélizumab |
| **sub-008** | N/A | None | None |
| **sub-009** | 0 | None | Switch from natalizumab to alemtuzumab |
| **sub-010** | N/A | None | None |
| **sub-011** | N/A | None | None |
| **sub-012** | 0 | None | None |
| **sub-013** | N/A | None | None |
| **sub-014** | 0.21 | 1 between T0 and T1/2 | Start Glatiramer acetate after the relapse |
| **sub-015** | N/A | None | Start ocrélizumab at T1/2 |
| **sub-016** | N/A | None | Start ocrélizumab at T1/2 |
| **sub-017** | 0.43 | 2 between T0 and T1 | Start Glatiramer acetate after first relapse |

**Supplementary data 4**: Line plots illustrating individual ARoC's for PD (left) and R1 (right) in NAWM. Each line corresponds to one subject. Dotted lines represent increasing rates.

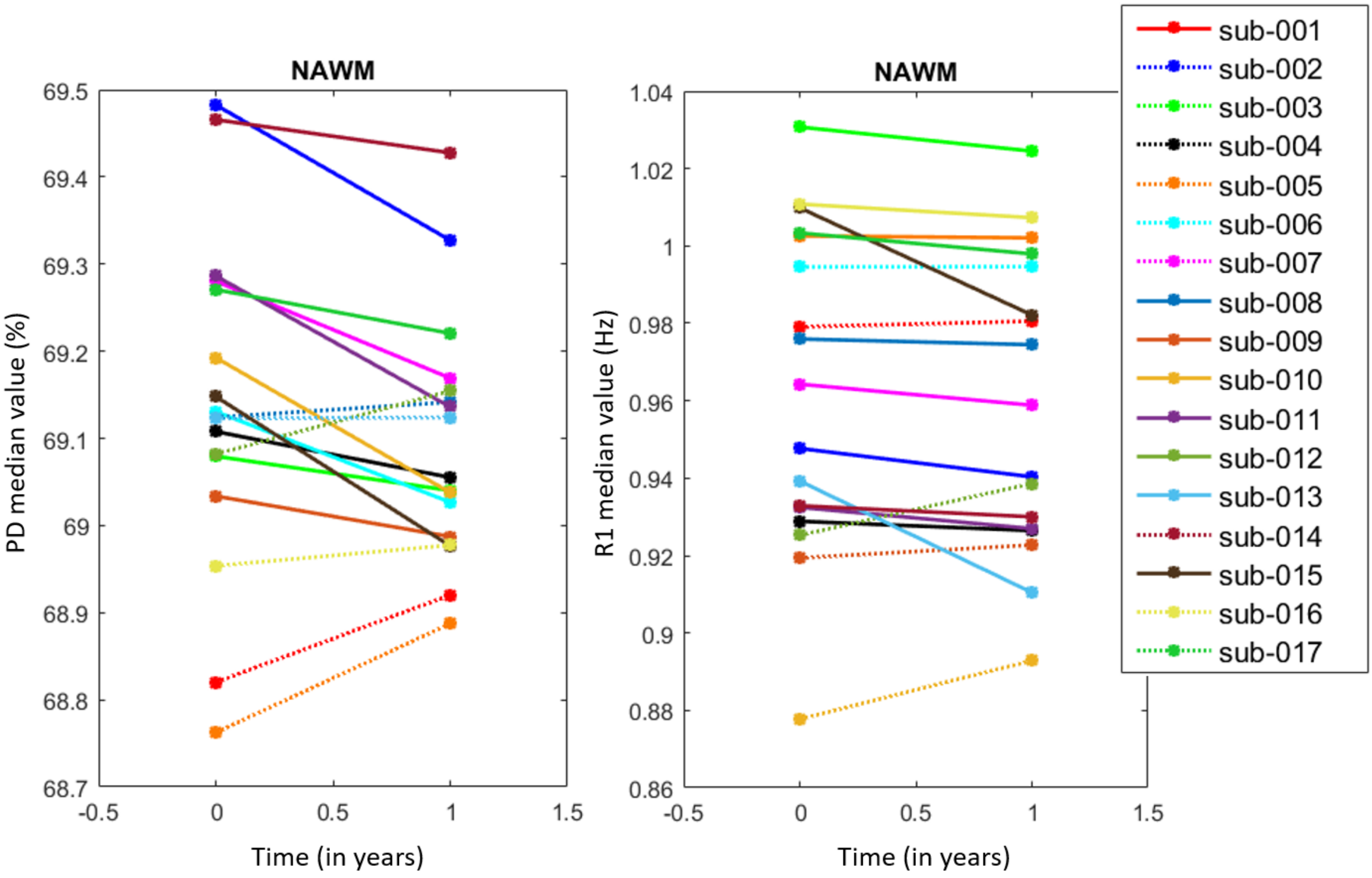

**Supplementary data 5:** Differences of lesion class Least Squares Means. First two columns correspond to tissue class labels (0 = NAWM, 1 = Later peripheral lesion, 2 = Initial peripheral lesion, 3 = FLAIR lesion).

### MTsat

| TISSUE_CLASS | _TISSUE_CLASS | Estimate | Standard Error | DF | t Value | Pr > \|t\| |
|---|---|---|---|---|---|---|
| 0 | 1 | 0.1761 | 0.06105 | 2956 | 2.88 | .0040 |
| 0 | 2 | 0.4153 | 0.06105 | 2956 | 6.80 | <.0001 |
| 0 | 3 | 0.4914 | 0.06105 | 2956 | 8.05 | <.0001 |
| 1 | 2 | 0.2393 | 0.04084 | 2427 | 5.86 | <.0001 |
| 1 | 3 | 0.3153 | 0.04084 | 2427 | 7.72 | <.0001 |
| 2 | 3 | 0.07604 | 0.04084 | 2427 | 1.86 | .0627 |

# PD

| TISSUE_CLASS | _TISSUE_CLASS | Estimate | Standard Error | DF | t Value | Pr > \|t\| |
|---|---|---|---|---|---|---|
| 0 | 1 | -0.03620 | 0.008791 | 4040 | -4.12 | <.0001 |
| 0 | 2 | -0.07043 | 0.008791 | 4040 | -8.01 | <.0001 |
| 0 | 3 | -0.09631 | 0.008791 | 4040 | -10.96 | <.0001 |
| 1 | 2 | -0.03422 | 0.005147 | 3395 | -6.65 | <.0001 |
| 1 | 3 | -0.06010 | 0.005147 | 3395 | -11.68 | <.0001 |
| 2 | 3 | -0.02588 | 0.005147 | 3395 | -5.03 | <.0001 |

# R1

| TISSUE_CLASS | _TISSUE_CLASS | Estimate | Standard Error | DF | t Value | Pr > \|t\| |
|---|---|---|---|---|---|---|
| 0 | 1 | 0.1004 | 0.03359 | 3265 | 2.99 | .0028 |
| 0 | 2 | 0.2187 | 0.03359 | 3265 | 6.51 | <.0001 |
| 0 | 3 | 0.2938 | 0.03359 | 3265 | 8.75 | <.0001 |
| 1 | 2 | 0.1183 | 0.02215 | 2718 | 5.34 | <.0001 |
| 1 | 3 | 0.1934 | 0.02215 | 2718 | 8.73 | <.0001 |
| 2 | 3 | 0.07516 | 0.02215 | 2718 | 3.39 | .0007 |

# R2*

| TISSUE_CLASS | _TISSUE_CLASS | Estimate | Standard Error | DF | t Value | Pr > \|t\| |
|---|---|---|---|---|---|---|
| 0 | 1 | 0.1043 | 0.03137 | 3927 | 3.32 | .0009 |
| 0 | 2 | 0.1672 | 0.03137 | 3927 | 5.33 | <.0001 |
| 0 | 3 | 0.2499 | 0.03137 | 3927 | 7.97 | <.0001 |
| 1 | 2 | 0.06288 | 0.01851 | 3236 | 3.40 | .0007 |
| 1 | 3 | 0.1456 | 0.01851 | 3236 | 7.86 | <.0001 |
| 2 | 3 | 0.08273 | 0.01851 | 3236 | 4.47 | <.0001 |
